# Antihypertensive medications and risk of Parkinson’s Disease: multi-cohort trial emulation and transcriptomic analyses

**DOI:** 10.1101/2025.05.20.25327943

**Authors:** Yuxuan Hu, Weiqiang Liu, Yujia Zhou, Miles Waits, Yiyong Zhao, Maria I. Olivero-Acosta, Katherine Sheng, Le Zhang, Hua Xu, Clemens R. Scherzer, Xianjun Dong

**Affiliations:** Stephen & Denise Adams Center for Parkinson’s Disease Research of Yale School of Medicine, New Haven, CT 06510, USA; Department of Neurology, Yale School of Medicine, New Haven, CT 06510, USA; Department of Neurology, Brigham and Women’s Hospital and Harvard Medical School, Boston, MA 02115, USA; Aligning Science Across Parkinson’s (ASAP) Collaborative Research Network, Chevy Chase, MD 20815, USA; Department of Biomedical Informatics and Data Science, Yale School of Medicine, New Haven, CT 06510, USA; Milton Academy, Milton, MA 02186, USA; Department of Neuroscience, Yale School of Medicine, New Haven, CT 06510, USA; Wu Tsai Institute, Yale University, New Haven, CT 06510, USA; Department of Genetics, Yale School of Medicine, New Haven, CT 06510, USA

**Author notes:** Correspondence should be addressed to: Xianjun Dong, PhD, Associate Professor, Departments of Neurology, Neuroscience, and Biomedical Informatics and Data Science Stephen & Denise Adams Center for Parkinson’s Disease Research, Yale School of Medicine 101 College Street, New Haven, CT 06510. These authors contributed equally (Yuxuan Hu, Weiqiang Liu).

**Keywords:** Parkinson’s disease, Target Trial Emulation, Multi-cohort, Antihypertensive Medications, Renin-angiotensin System

## Abstract

**Background:** No known treatments prevent Parkinson’s disease (PD) onset or delay its progression. Prior observational studies of antihypertensive medications and PD risk have produced inconsistent findings and may be susceptible to confounding and reverse causation.

**Methods:** We applied a multi-stage analytical framework integrating medication-wide screening, independent cohort replication, target trial emulation, and exploratory transcriptomic analyses. Medication-wide association screening was conducted in a large-scale electronic health record (EHR)-linked biobank using logistic regression, with replication in independent cohorts. For candidate drug classes, we emulated a target trial among individuals with primary hypertension initiating first-line antihypertensive therapy in the Merative™ MarketScan^®^ Medicare Database. Incident PD was the primary outcome, and associations were estimated using inverse-probability-of-treatment-weighted Cox proportional hazards models. Exploratory transcriptomic analyses integrated single-cell drug-perturbation data with PD brain expression datasets.

**Findings:** Across medication-wide screening and independent cohort replication, losartan (an ARB) and amlodipine (a CCB) were consistently associated with lower odds of PD. The target trial emulation included 269,617 individuals with primary hypertension (25,965 ARB initiators; 36,873 CCB initiators; and 206,779 initiators of other first-line antihypertensives). ARB initiation was associated with a lower hazard of incident PD than initiation of other first-line antihypertensives (adjusted HR [aHR] 0.70, 95% CI 0.59–0.83); the corresponding association for CCB initiation was smaller (aHR 0.88, 95% CI 0.77–0.99). The ARB association was generally consistent across sensitivity analyses addressing alternative exposure definitions, measured prodromal PD characteristics, and comparator choice, but was attenuated at a 5-year PD outcome lag. ARB initiation was also associated with a lower PD risk than ACE inhibitor initiation (aHR 0.81, 95% CI 0.67–0.97). Exploratory transcriptomic analyses showed directional opposition between PD-associated molecular signatures and transcriptional changes after sacubitril/valsartan exposure.

**Interpretation:** ARBs initiation was associated with a lower risk of incident PD than initiation of other first-line antihypertensive medications. The attenuation observed for 5-year lag time leaves room for alternative explanations including the influence of prodromal disease on ARB use, delayed ascertainment, and other possible sources of bias. These findings warrant prospective evaluation but do not endorse ARB use for the clinical prevention of PD.

## Introduction

Parkinson’s disease (PD) is a common neurodegenerative disease characterised by motor and non-motor manifestations and is estimated to affect 13 million people by 2040.^1,2^ No disease-modifying therapy has been proven to prevent PD onset or delay its progression. Hypertension is highly prevalent among older adults, who are also at increased risk of PD, and vascular, neuroinflammatory, and neurodegenerative processes may intersect in PD pathogenesis.^3,4^ Antihypertensive medications, including angiotensin II receptor blockers (ARBs) and calcium channel blockers (CCBs), have been hypothesized to have neuroprotective effects through modulation of cerebral perfusion, neuroinflammation, oxidative stress, and neuronal signalling pathways.^5–8^

However, epidemiological evidence linking antihypertensive medications to PD remains inconsistent.^9–15^ Although several observational studies and meta-analyses have reported inverse associations between CCB use and PD incidence, other prospective studies have found no significant association.^16,17^ Evidence for renin–angiotensin system inhibitors has also been mixed, with pooled estimates from cohort studies showing no statistically significant associations for ARBs or angiotensin-converting enzyme inhibitors (ACEIs).^18^ More recently, a prospective study of French women found no significant association between overall CCB use and PD after incorporating a five-year exposure lag, while reporting higher PD incidence for certain individual agents.^19^ Furthermore, a drug-target Mendelian randomization study found little evidence for a causal association between genetically proxied antihypertensive drug-target perturbation on PD risk or age at onset.^15^ In contrast, Jo et al. reported a lower PD risk among users of blood-brain barrier (BBB)-crossing ARBs, but not non-BBB-crossing ARBs, in a Korean population with ischemic heart disease, suggesting potential heterogeneity within the ARB class.^12^ Although genetic proxies for drug-target inhibition do not necessarily reproduce the effects of pharmacological treatment, these findings underscore the uncertainty surrounding the potential neuroprotective effects of antihypertensive medications. Together, the conflicting evidence highlights the need for studies employing rigorous pharmacoepidemiological designs, clinically relevant active comparators, and complementary approaches to assess biological plausibility.

To address these uncertainties, we conducted a multi-stage observational study integrating drug-wide association screening in the Mass General Brigham (MGB) Biobank dataset,^20^ independent replication in two Accelerating Medicines Partnership Parkinson’s Disease cohorts (Parkinson’s Progression Markers Initiative [PPMI] and Parkinson’s Disease Biomarkers Program [PDBP]),^21^ and target trial emulation (TTE) using the Merative™ MarketScan® Medicare Database among individuals with hypertension, with over 10 years of follow-up period. Medication-wide screening and independent replication allowed us to identify and evaluate associations across datasets, while TTE provided a framework for comparing treatment initiation strategies using clinically relevant active comparators when randomised trials are unavailable.^22^ Recognising that observational analyses remain susceptible to residual confounding, confounding by indication, reverse causation, treatment-selection bias, and exposure or outcome misclassification, we performed sensitivity and subgroup analyses addressing potential reverse causation and alternative exposure, outcome, and comparator definitions, including exploratory analysis of BBB-crossing versus non-BBB-crossing ARBs and individual ARB agents. Finally, exploratory transcriptomic analyses were incorporated to provide biological context for the epidemiological findings, rather than mechanistic validation.

## Methods

### Ethics

The Mass General Brigham (MGB) Biobank component of this study was conducted under approval from the [Mass General Brigham/Research Patient Data Registry] (IRB no. [2005P001191]). The Merative™ MarketScan® Medicare Database provided de-identified administrative claims data accessed under an institutional data use agreement. The PPMI and PDBP analyses used de-identified cohort data obtained through their respective approved data-access procedures and data use agreements (DUA). No participants were contacted or enrolled by the investigators for these analyses. The transcriptomic analyses used previously published cell line and human post-mortem brain RNAseq data, therefore non-human subject research. No additional participants were recruited for the present transcriptomic analyses.

### Study design and data sources

Drug-wide screening was performed using the MGB Biobank, a large EHR-linked research cohort at Mass General Brigham, as the discovery cohort. Replication analyses used two Accelerating Medicines Partnership Parkinson’s Disease (AMP-PD) cohorts, PPMI and PDBP. We emulated target trials using claims from the Merative™ MarketScan^®^ Medicare Database (MDCR), which includes Medicare-eligible retirees with employer-sponsored Medicare Supplemental or Medicare Advantage coverage. Among individuals with hypertension, we used an active-comparator, new-user design to compare initiation of first-line antihypertensive therapy.^23,24^ The TTE design is shown in **Supplementary Figure 1**.

Transcriptomic analyses incorporated single-cell perturbation data from the Tahoe-100M dataset and a single-cell RNA sequencing dataset from PD brain tissue.^25,26^ All datasets were de-identified and analysed under the applicable institutional review board approvals or data use agreements. Reporting followed the Strengthening the Reporting of Observational Studies in Epidemiology (STROBE) and the Transparent Reporting of Observational Studies Emulating a Target Trial (TARGET) guidelines.^27^ The study population and analytical role for the datasets are summarized **Supplementary Table 1**.

## Study population

### Drug-Wide Screening and Replication

To identify PD cases comprehensively, PD cases in the MGB Biobank were ascertained using ICD codes (ICD-9-CM: 332, 332.0; ICD-10-CM: G20, G20.A1, G20.A2, G20.B1, G20.B2, G20.C) and hospital-specific codes (LMR: LPA312, LPA883; ONCALL: WHJM1; DSM4: 332..3). Index dates were defined as the first recorded qualifying PD diagnosis for cases and last recorded encounter in the EHR system for controls. Individuals with a recorded diagnosis of dementia on or before the PD index date were excluded from the primarily identified PD cases. Each eligible PD case was matched to up to two controls using nearest-neighbour matching based on propensity score estimated from age, sex, and race, with a calliper of 0.1. Detailed inclusion and exclusion criteria are provided in the **Supplementary Methods**. Replication cohorts used standardized AMP-PD diagnostic criteria, with enrolment date as the index date. Demographic, diagnostic, and prescription data were harmonized across cohorts, and consistent inclusion and exclusion criteria were applied to ensure comparability (**Supplementary Methods**). **Target Trial Emulation**: The TTE cohort included MDCR beneficiaries aged ≥ 18 years with primary hypertension initiating first-line antihypertensive medications between January 2009 and September 2019. Hypertension was identified using ICD-9-CM code 401.X or ICD-10-CM code I10. The dispensing date of the first qualifying antihypertensive prescription was defined as the index date. Eligible individuals were required to have at least one year of continuous insurance enrolment before the index date and no use of any first-line antihypertensive medication during this baseline period, consistent with a new-user design. They were also required to have filled at least one prescription of first-line antihypertensive agents, including ARB, CCB, or other first-line antihypertensive medication (angiotensin-converting enzyme inhibitor [ACEI], β-blocker [BB], or thiazide).^28^ Patients with a prior diagnosis of PD (ICD-9-CM: 332.0 or ICD-10-CM: G20) or use of anti-Parkinson disease medications (**Supplementary Table 2**) before the index date were excluded.^29,30^ To identify individuals with a stable initial treatment strategy, we additionally applied a 90-day treatment-stability window after the index date. Individuals who developed PD (based on ICD code above) or switched to or added another study antihypertensive class during this window were also excluded from the primary analysis. Follow-up for incident PD began on day 91 after treatment initiation. The full protocol with additional eligibility criteria specified in **Supplementary Table 3**.

### Exposure definition

Medication exposure was defined using prescription records mapped to standardized drug identifiers (**Supplementary Methods** and **Supplementary Table 2**). In the drug-wide association screening, medication exposure was defined as at least one recorded prescription prior to the index date. Analyses were restricted to the medications prescribed to at least 10 individuals. Drug classes were identified using Anatomical Therapeutic Chemical (ATC) codes. In the TTE analysis, treatment groups were defined according to the first-line antihypertensive class initiated on the index date. A 90-day treatment-stability window was applied after initiation, as described above, and the primary outcome follow-up began on day 91.

### Outcome assessment

In the drug-wide screening and replication analyses, the outcome was PD cases status as defined according to the diagnostic criteria described in the population design. In the primary TTE analysis, follow-up for incident PD began on day 91 after treatment initiation and continued for up to 10 years until the first qualifying PD diagnosis, switching to or addition of another study antihypertensive class, loss of insurance eligibility, or the end of the study period (September 30, 2024), whichever occurred first^31,32^. Additional censoring at treatment discontinuation was evaluated in a sensitivity analysis, with discontinuation defined as 90 days after the last observed prescription for the initially assigned treatment strategy. Secondary analyses evaluated 5-year and all-available follow-up horizons.

### Covariates

In the drug-wide association analyses, demographic characteristics (age, sex, race) were applied as covariates, and models were also adjusted for source (PPMI or PDBP) in the replication cohort. In the TTE, we included patient demographics (age, sex, and year of enrolment), health care utilization (number of outpatient visits and hospitalizations), and 10 comorbidities (congestive heart failure, cardiac arrhythmias, valvular disease, peripheral vascular disease, other neurological disorders, chronic pulmonary disease, diabetes uncomplicated, diabetes complicated, renal failure, and depression) occurring within the baseline period and classified as present if patient had at least one diagnosis code using Elixhauser Comorbidity Index (ECI) (**Supplementary Table 4**).^30,31,33^ To address potential confounding factors related to prodromal PD and treatment selection, an expanded propensity score model additionally included orthostatic hypotension, syncope, falls, cerebrovascular disease, tremor, constipation, REM sleep behaviour disorder, olfactory impairment, dopamine-blocking medication use, and overall baseline medication burden (**Supplementary Table 5**).

### Statistical analysis

Medication-wide screening used logistic regression to estimate associations between individual medications and PD risk, adjusting for age, sex, and race (**Supplementary Table 4**). In the replication dataset, models were also adjusted for source (PPMI or PDBP). Specifically, a logistic regression model was employed to estimate the association between the drug exposure and the risk of developing PD for each drug. The odds ratio (OR) and corresponding 95% confidence interval (CI) were calculated, with statistical significance assessed by *P* values. False discovery rate (FDR) correction (Benjamini-Hochberg) was applied. Drugs showing an association with reduced PD risk in the discovery analysis were required to have an OR < 1 and an FDR < 0.05, whereas drugs in the replication analysis were required to have an OR < 1 and a nominal *P* value < 0.05. Replication was assessed at the generic-drug level, with drug names harmonized across the two analyses. A final list of replicated drugs was defined as those meeting these criteria in both cohorts and sharing the same harmonized generic drug identity. Substantial sensitivity analyses were undertaken to assess the robustness of the drug-wide screening results to PD cases definition, potential delay in code-based PD ascertainment, reverse causation, and changes in antihypertensive treatment preceding PD diagnosis. Specifically, PD definition sensitivity analyses included restriction to cases with PD diagnoses recorded in neurology clinics as a proxy to neurologist-confirmed diagnosis, repeated PD diagnoses on at least two distinct dates, recorded levodopa or dopamine-agonist treatment, or exclusion of individuals with post-index secondary parkinsonism. Since the first recorded PD code might occur after disease onset or treatment initiation, we quantified the timing of dopaminergic treatment relative to the first qualifying PD code and implemented sensitivity analyses after excluding patients with more than 180-day pre-code dopaminergic treatment and after shifting the index date 1, 2, and 5 years earlier. We also examined antihypertensive treatment-intensity trajectories during the five years preceding the index date to assess potential de-prescribing during the prodromal period. Additional analyses varied the pre-index exposure window and exposure definition and adjusted for concurrent antihypertensive treatment and overall medication burden (**Supplementary Methods**).

In the TTE, we applied inverse probability of treatment weighting (IPTW) based on propensity scores to estimate the average treatment effect (ATE), aiming to maintain statistical power and precision, especially when dealing with rare outcomes.^32,34^ Propensity scores for treatment assignment were estimated separately for each pairwise comparison using a multivariable logistic regression model that included all the baseline covariates. The full model coefficients are reported in **Supplementary Table 6**. Stabilized IPTW weights were subsequently applied to create weighted cohorts and balance baseline characteristics across treatment groups. We also implemented the analyses using an expanded propensity score model with additional covariates related to treatment selection and PD risk. Covariate balance before and after weighting was assessed using absolute standardized mean differences (SMDs), with values less than 0.1 indicating adequate balance.^35^ Where residual covariate imbalance remained after weighting, an additional sensitivity analysis further adjusted the weighted Cox model for these characteristics. IPTW weights were trimmed at the 1st and 99th percentiles.^35,36^ IPTW-weighted Cox proportional hazards models with robust sandwich variance estimation were used to estimate the association between initiation of ARBs or CCBs and PD risk, with other first-line antihypertensive agents serving as the active comparator group. Adjusted hazard ratios (aHRs) and corresponding 95% CIs were reported. Crude and IPTW-weighted cumulative PD risks were estimated at 5 and 10 years, and corresponding absolute risk differences between treatment groups were calculated. *P* values were two-sided, and *P* < 0.05 was considered statistically significant. We conducted the following subgroup analyses: (1) age (<50, 50-64, or ≥65); (2) sex (female or male). A series of sensitivity analyses evaluated alternative design and analytic assumptions, including (1) using propensity score-matched Cox model with a ratio of 1:1; (2) using an unweighted Cox model; (3) excluding BBs initiators in the other first-line antihypertensive group; (4) initiating the follow-up period from the index date to capture outcome events that occurred early; (5) extending treatment-stability window from 90 days to 180 days to examine sensitivity to the definition of stable initial treatment; (6) redefining the outcome using at least two outpatient diagnoses of PD during follow-up; (7) altering the ICD-10-CM code for PD outcome definition to G20 and G20.X; (8) using an intention-to-treat strategy; and (9) adjusting for unbalanced covariates; (10) using expanded PS model with additional covariates; and (11) additional censoring at treatment discontinuation. To address potential reverse causation related to long prodromal phase of PD or delayed PD ascertainment, we additionally conducted 1-, 2-, and 5-year PD outcome lag analyses in which the start of PD risk follow-up was delayed by the corresponding interval after treatment initiation. Patients who developed PD or were censored before the respective risk-entry time were excluded. To evaluate the influence of comparator choice, class-specific active-comparator analyses compared ARB initiators separately with ACEI and CCB initiators. Propensity scores and IPTW weights were re-estimated independently for each comparison using both the primary and expanded covariate sets. Exploratory analyses further evaluated potential heterogeneity according to BBB penetration (BBB-crossing ARBs: valsartan, telmisartan, fimasartan, candesartan, azilsartan; non-BBB-crossing ARBs: irbesartan, losartan, olmesartan, eprosartan)^12^ and individual ARB agents. The E-value was calculated as a quantitative sensitivity analysis to assess the robustness of the observed association to potential unmeasured confounding.^37,38^ The data analyses for this study were conducted from November 2023 to December 2025.

### Transcriptomic analysis

Drug-induced transcriptional signatures were derived from the Tahoe-100M dataset by comparing sacubitril/valsartan-treated A-172 astrocyte-like cells (5 μM) with matched DMSO-treated controls. Because the perturbation was the combination product, the resulting transcriptional signature cannot be attributed specifically to valsartan. PD-associated transcriptional signatures were obtained from published prefrontal cortical single-nucleus RNA-seq datasets, and pseudobulk profiles were generated for differential expression analysis, with age, sex, and post-mortem interval adjusted. Gene set enrichment analysis (GSEA) was used to compare pathway-level changes in the drug-perturbation and PD-associated signatures using Gene Ontology and KEGG gene sets. Differential expression results were corrected for multiple testing using the Benjamini-Hochberg method. Genes and pathways with an adjusted *P* < 0.05 were considered statistically significant. Among significant genes and pathways, directional concordance was assessed descriptively according to the direction of the corresponding effect estimates (log2 fold change [log2FC] for genes and normalized enrichment score [NES] for pathways), with opposite signs indicating potential reversal patterns between drug perturbation and PD-associated transcriptional signatures (see **Supplementary Methods** for details).

## Results

### Study population

The drug-wide association screening included 87,359 participants from the MGB Biobank, of whom 48,064 met eligibility criteria (16,797 PD cases; 31,267 controls; **Supplementary Table 7**). Among PD cases, 94.0% were older than 60 years, 58.7% were male, and 87.2% were recorded as White. Among the 16,797 PD cases, 70.8% had PD diagnoses recorded on at least two distinct dates, 72.5% had an outpatient record at the first qualifying PD diagnosis date, and 57.8% had at least one PD diagnosis recorded in a validated neurology clinic. The replication cohort included 2,351 PD cases with similar demographic characteristics (**Supplementary Figure 2**). Assessment of diagnostic timing showed that 3,731 (22.2%) of 16,797 PD cases had a recorded prescription for levodopa or a dopamine agonist before their first qualifying PD code; among these patients, treatment preceded the first PD code by a median of 205 days (interquartile range [IQR] 49-776.5), indicating that code-based ascertainment could occur after treatment initiation in a subset of patients (**Supplementary Results**). In contrast, antihypertensive treatment trajectories during the five years preceding the index date showed no evidence of a general decline in treatment intensity among PD cases relative to controls (**Supplementary Results and Supplementary Figure 3**).

In the TTE, 269,617 individuals with primary hypertension initiating first-line antihypertensive medications between 2009 and 2019 were included (25,965 ARB initiators; 36,873 CCB initiators; 206,779 other first-line antihypertensive initiators). Baseline characteristics are shown in **Table 1**. Among the 2,766 incident PD events included in the primary analysis (**Table 2**), 76.0% had PD diagnoses recorded on at least two distinct dates, and the first qualifying PD event was identified in an outpatient setting in 90.7%, an inpatient setting in 9.1%, and both settings in 0.2%. Compared with initiators of other first-line antihypertensives, ARB initiators were younger and had lower prevalence of congestive heart failure and cardiac arrhythmias, whereas CCB initiators were older and had a higher prevalence of chronic pulmonary disease. IPTW improved covariate balance in both comparisons. Detailed pre- and post-weighting baseline characteristics are provided in **Supplementary Table 8**. All measured baseline characteristics achieved an absolute SMD below 0.10 in the CCB comparison, whereas residual imbalance remained for hospitalizations, congestive heart failure, cardiac arrhythmias in the ARB comparison (**Table 1**). Similar to MGB Biobank, diagnostic-timing analyses in MDCR showed that levodopa or dopamine agonist use preceded the first qualifying PD code in a subset of patients (570 [20%] of 2,845 PD cases, median 95.5 days [IQR 38-329]), indicating that code-based PD ascertainment may lag treatment initiation.

**Table 1.**
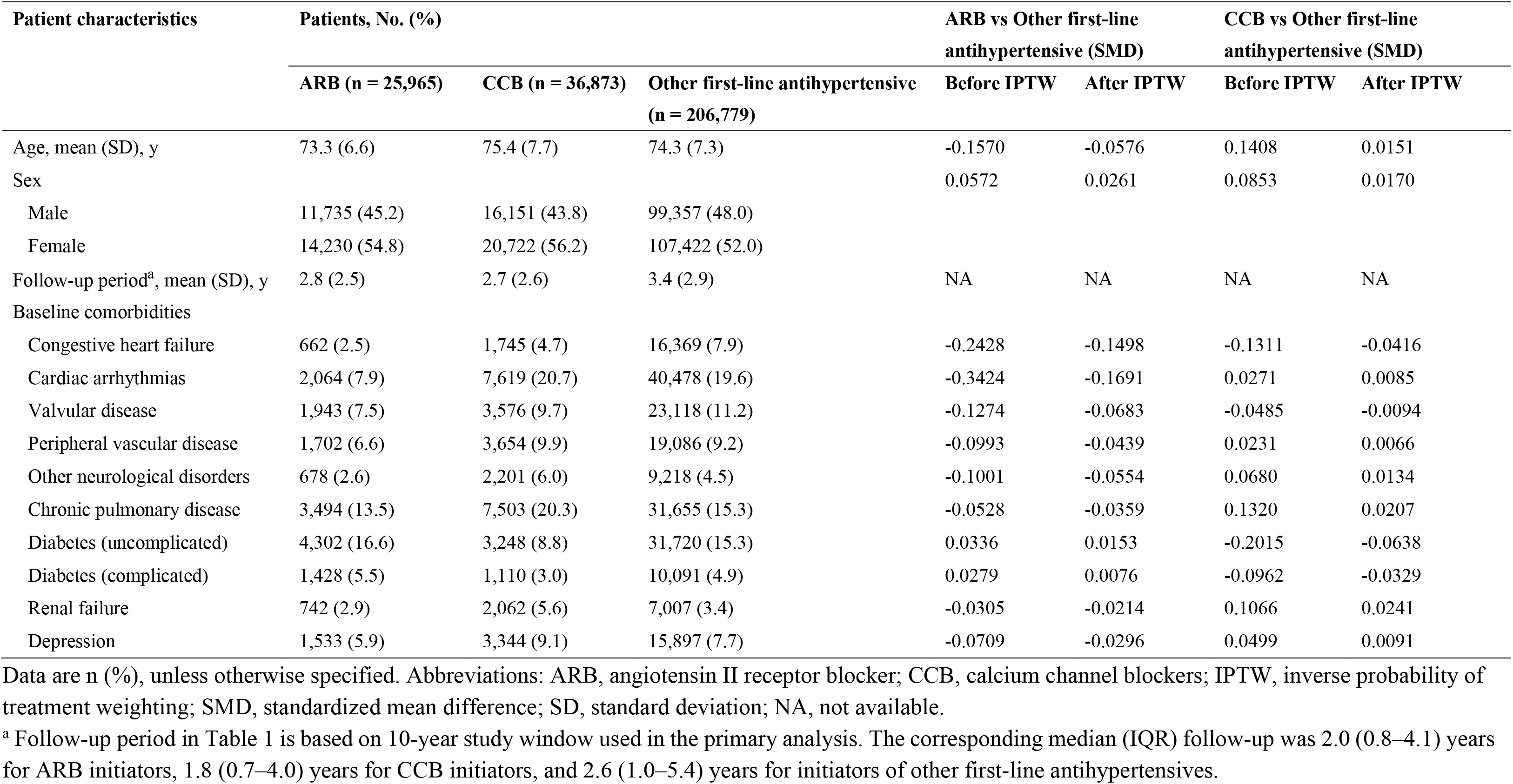
Baseline characteristics of the selected patients in target trial emulation.

**Table 2.** Associations between angiotensin II receptor blockers or calcium channel blockers and incidence of Parkinson’s disease in MarketScan Medicare

| Outcomes | ARB vs Other first-line antihypertensives |  |  |  |  | CCB vs Other first-line antihypertensives |  |  |  |  |
| --- | --- | --- | --- | --- | --- | --- | --- | --- | --- | --- |
|  | No. of PD event, percentage (%) |  | aHR (95% CI) | Absolute risk differences |  | No. of PD event, percentage (%) |  | aHR (95% CI) | Absolute risk differences |  |
|  |  |  |  | Crude | Adjusted |  |  |  | Crude | Adjusted |
|  | ARB (n = 25,965) | Other first-line antihypertensives (n = 206,779) |  |  |  | CCB (n= 36,873) | Other first-line antihypertensives (n = 206,779) |  |  |  |
| 10-year risk | 153 (0.59) | 2,322 (1.12) | 0.70 (0.59–0.83) | 0.53% | 0.54% | 291 (0.79) | 2,322 (1.12) | 0.88 (0.77–0.99) | 0.33% | 0.79% |
| 5-year risk | 116 (0.45) | 1,782 (0.86) | 0.63 (0.52–0.77) | 0.42% | 0.56% | 246 (0.67) | 1,782 (0.86) | 0.89 (0.77–1.02) | 0.20% | 0.30% |
| Subgroup analyses |  |  |  |  |  |  |  |  |  |  |
| Age |  |  |  |  |  |  |  |  |  |  |
| <50 | 0/26 | 0/169 | NA |  |  | 0/29 | 0/169 | NA |  |  |
| 50-64 | 5/523 | 41/4,426 | 1.55 (0.58–4.14) |  |  | 1/747 | 41/4,426 | 0.16 (0.02–1.19) |  |  |
| ≥65 | 148/25,416 | 2,281/202,184 | 0.69 (0.58–0.82) |  |  | 290/36,097 | 2,281/202,184 | 0.89 (0.79–1.01) |  |  |
| Sex |  |  |  |  |  |  |  |  |  |  |
| Male | 93/11,735 | 1,465/99,357 | 0.75 (0.60–0.94) |  |  | 158/16,151 | 1,465/99,357 | 0.87 (0.74–1.04) |  |  |
| Female | 60/14,230 | 857/107,422 | 0.65 (0.50–0.86) |  |  | 133/20,722 | 857/107,422 | 0.92 (0.76–1.11) |  |  |

Adjusted absolute risk differences were estimated from the IPTW-weighted cumulative risks at the specified time points (5-year and 10-year). Abbreviations: ARB, angiotensin II receptor blockers; CCB, calcium channel blockers; PD, Parkinson’s disease; aHR, adjusted hazard ratio; CI, confidence interval; NA, not available.

### Medication-wide screening and replication analyses identified antihypertensive drugs associated with lower PD risk

Among a total of 1,601 medications in discovery and 305 medications in replication, five medications were consistently associated with lower PD risk in both discovery and replication analyses (**Figure 1A**), including two antihypertensive agents: losartan (ARB) and amlodipine (CCB). In the MGB Biobank, losartan use was associated with lower risk of PD (odds ratio [OR] 0.88, 95% confidence interval [CI], 0.82–0.94), with a stronger association observed in the replication cohorts (OR 0.67; 95% CI 0.51–0.88). Amlodipine showed similar associations in discovery (OR 0.89, 95% CI 0.85–0.94) and replication analyses (OR 0.73, 95% CI 0.57–0.93). Ten other antihypertensive medications were significantly associated with reduced PD risk in discovery cohort alone (e.g. they are either not included or not significant in replication cohort), including two aldosterone antagonists (spironolactone and eplerenone), one ARB (valsartan), one CCB (nicardipine), five β-blockers (metoprolol, esmolol, labetalol, carvedilol, and landiolol), and one thiazide (chlorothiazide). More details on the associations between antihypertensive medications and PD risk in the discovery analysis are visualized in **Supplementary Figure 4**. The associations of losartan or amlodipine with reduced PD risk were generally maintained across sensitivity analyses using more stringent PD cases and alternative temporal and exposure definitions (**Figure 1B**). Restriction to neurologist-supported PD yielded ORs of 0.88 (95% CI 0.81–0.96) for losartan and 0.87 (95% CI 0.81–0.93) for amlodipine. Importantly, the associations persisted when the index date was shifted progressively earlier to reduce potential reverse causation. The estimates were also broadly consistent after adjustment for concurrent antihypertensive classes and overall medication burden, although estimates were attenuated in some analyses using more stringent medication-exposure definitions (**Figure 1B**).

**Figure 1.**
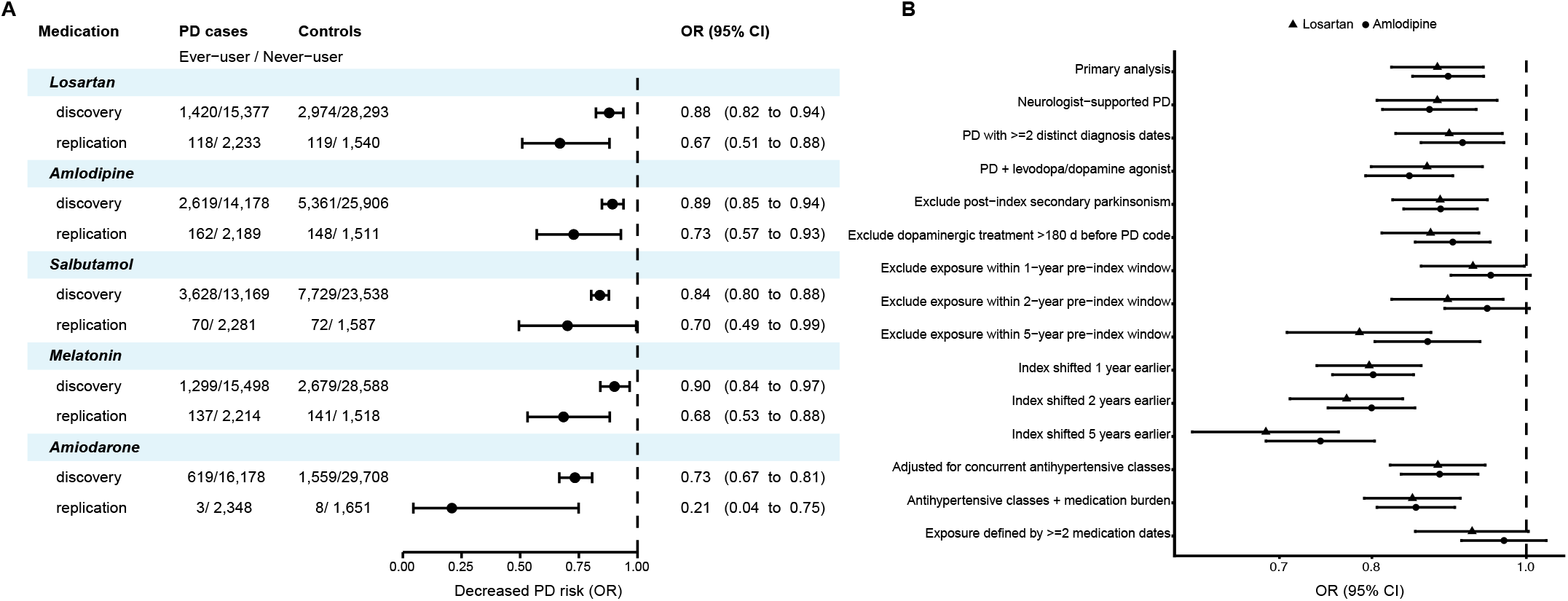
Two antihypertensive medications significantly associated with reduced Parkinson’s disease risk. (A) Forest plot of five drugs that were significantly associated with reduced PD risk. MGB Biobank was used as the discovery cohort for drug-wide screening and two AMP-PD cohorts, PPMI and PDBP, were used as the discovery cohorts. (**B**) Results of sensitivity analyses in the drug-wide screening analysis. Abbreviations: MGB, Mass General Brigham; PPMI, Parkinson’s Progression Markers Initiative; PDBP, Parkinson’s Disease Biomarkers Program; PD, Parkinson’s disease; AMP-PD, Accelerating Medicines Partnership Parkinson’s Disease; OR, odds ratio; CI, confidence interval.

### Association between antihypertensive initiation and incident PD in target trial emulation

Given the consistent association of antihypertensive agents with reduced PD risk in the drug-wide association and replication analyses, we next evaluated these findings in a TTE study among individuals with hypertension using MDCR data. Participant selection is summarized in **Figure 2A**. After applying baseline eligibility criteria and the 90-day treatment-stability window, 269,617 individuals with primary hypertension were included (25,965 ARB initiators, 36,873 CCB initiators, and 206,779 initiators of other first-line antihypertensives). In the primary analysis, follow-up for incident PD began on day 91 after treatment initiation.

**Figure 2.**
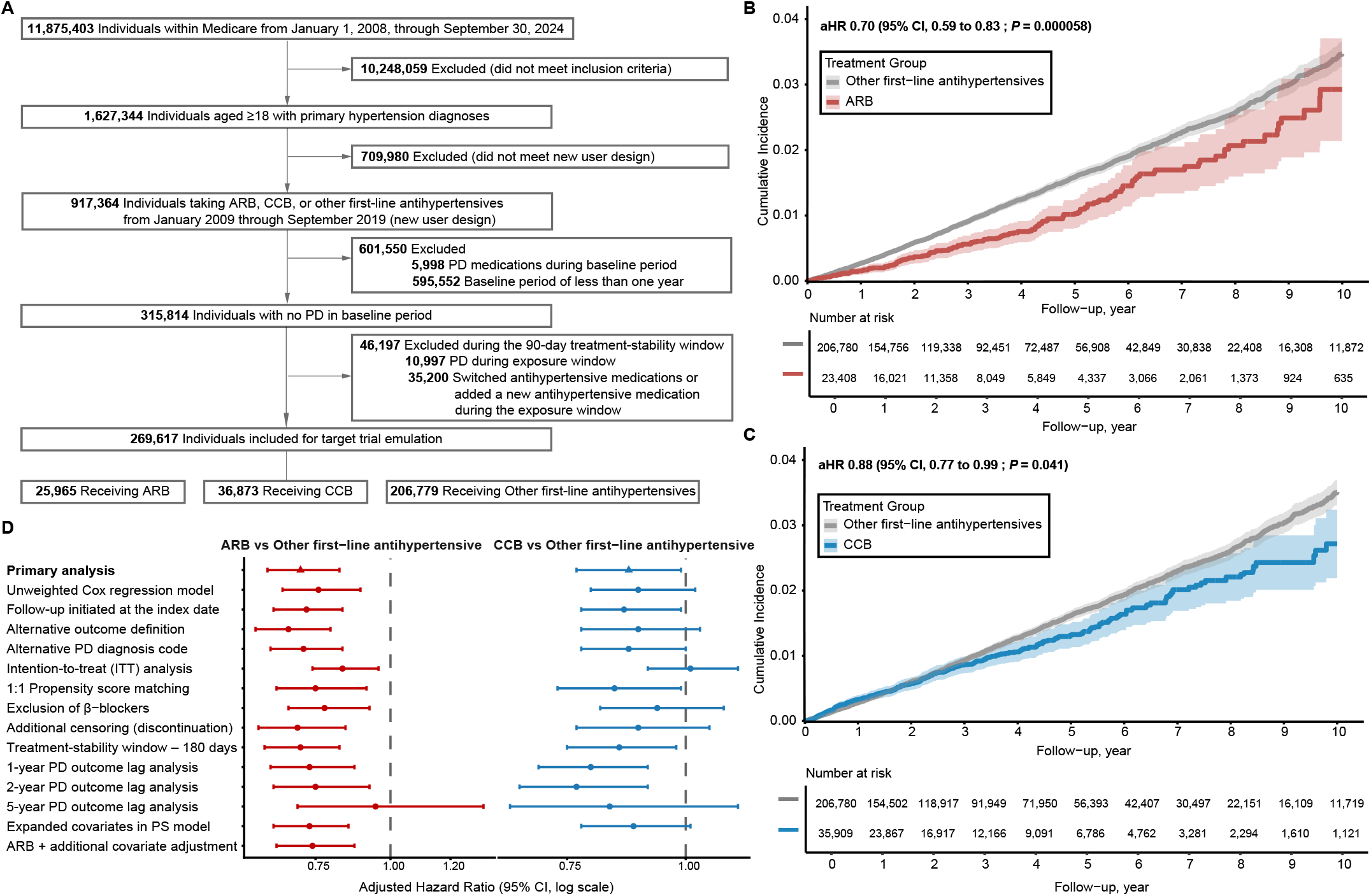
Associations between antihypertensive initiation and 10-year risk of Parkinson’s disease in target trial emulation. (**A**) Flow diagram of participant selection in target trial emulation. The inclusion criteria and new-user design criteria are presented in **Supplementary Table 3**. Among individuals with primary hypertension, 490,218 were excluded because none first-line antihypertensive treatment was identified, 127,172 because treatment initiation occurred too late to allow at least 5 years of potential administrative follow-up, 87,488 because of simultaneous initiation of two or more treatment groups, and 5,102 because of initiation with ARB+CCB fixed-dose combination. (**B**) Inverse probability of treatment weighting (IPTW)-adjusted cumulative incidence of Parkinson’s disease comparing ARB initiators with initiators of other first-line antihypertensives. (**C**) IPTW-adjusted cumulative incidence of Parkinson’s disease comparing CCB initiators with initiators of other first-line antihypertensives. Shaded areas represent 95% CIs. Numbers at risk are shown below the x-axis. (**D**) Forest plot of sensitivity analyses for target trial emulation. Abbreviations: ARB, angiotensin II receptor blocker; CCB, calcium channel blocker; PD, Parkinson’s disease; aHR, adjusted hazard ratio; CI, confidence interval; ITT, intention-to-treat.

ARB initiation was associated with a significantly lower hazard of incident PD over 10 years compared with initiation of other first-line antihypertensives (aHR 0.70, 95% CI 0.59–0.83; adjusted absolute risk difference 0.54%) (**Table 2**). The estimate was stronger in the 5-year analysis (aHR 0.63, 95% CI 0.52–0.77; adjusted absolute risk difference 0.56%). CCB initiation was associated with a smaller difference in PD hazard over 10 years (aHR 0.88, 95% CI 0.77–0.99; adjusted absolute risk difference 0.79%), with attenuated estimates in the 5-year analysis (aHR 0.89, 95% CI 0.77–1.02; adjusted absolute risk difference 0.30%). Similar associations were also observed when follow-up window was extended over all-available horizon (**Supplementary Figure 5**). IPTW-adjusted Kaplan-Meier curves showed early and persistent separation between treatment groups throughout the 10-year (**Figure 2B-C**). The E-value was 2.21 for ARBs and 1.53 for CCBs, meaning that, conditional on the measured covariates, an unmeasured confounder associated with both treatment and PD by risk ratios of at least these magnitudes could explain the corresponding point estimates.

### Subgroup and Sensitivity Analyses

The association between ARB initiation and PD was generally consistent across prespecified subgroups, with no strong evidence of effect modification by age groups and sex (**Table 2**). Results were also similar in sensitivity analyses using alternative exposure definitions and an expanded propensity-score (PS) model (**Figure 2D, Supplementary Tables 9–12**). An analysis beginning outcome follow-up on the index date, rather than after the 90-day treatment-stability window, was also conducted to assess sensitivity to delayed entry into follow-up (aHR 0.72, 95% CI 0.61–0.84).

In analyses incorporating progressively longer lags between antihypertensive initiation and PD ascertainment, the association was similar with 1- and 2-year lags but was substantially attenuated with a 5-year lag (aHR 0.95, 95% CI 0.69–1.31; **Figure 2D**). This attenuation with a longer lag suggests that reverse causation related to prodromal PD cannot be excluded.

In additional active-comparator analyses, ARB initiation was associated with a lower hazard of PD compared with ACEI initiation (aHR 0.81, 95% CI 0.67–0.97), with a similar estimate using the expanded PS model (aHR 0.82, 95% CI 0.68–0.99), whereas no significant difference was observed compared with CCB initiation (**Supplementary Table 10**). Covariates were adequately balanced after weighting in these pairwise comparisons (**Supplementary Figure 6**). Exploratory analyses according to BBB penetration and individual ARB agents are reported in **Supplementary Tables 11–12**. More method and result details for sensitive analyses can be found in Supplementary results.

### Transcriptomic Analyses Supporting Biological Plausibility

Single-cell perturbation data from sacubitril/valsartan-treated A-172 astrocyte-like cells were compared with PD-associated gene expression profiles of astrocytes and microglia in human post-mortem brain. Four genes (*ZFP36, FAM162A, MT1X,* and *IL7R*) were significantly differentially expressed in both datasets, with opposite directions of effect between PD and sacubitril/valsartan perturbation (e.g., log2 fold changes 3.98 and -0.10, adjusted *P* = 5.78 × 10^-3^ and 6.10 × 10^-4^ for *ZFP36* in PD astrocytes and drug perturbation, respectively; **Figure 3A**). They are known regulators of inflammatory signalling (*ZFP36* and *IL7R*) and oxidative stress (*FAM162A* and *MT1X*). Pathway-level analyses also identified selected inflammatory, cellular-stress, proteostasis, and neurodegeneration-related pathways with opposing enrichment directions (e.g., NES 1.70 and -1.96, adjusted *P* = 2.91× 10^-4^ and 2.29 × 10^-4^ for Parkinson’s disease pathway in PD and drug perturbation, respectively; **Figure 3B-D; Supplementary Figure 7**). Because the perturbation used sacubitril/valsartan in a transformed astrocyte-like cell line and the disease signatures were derived from prefrontal cortex, these findings are exploratory and do not establish an ARB-specific mechanism.

**Figure 3.**
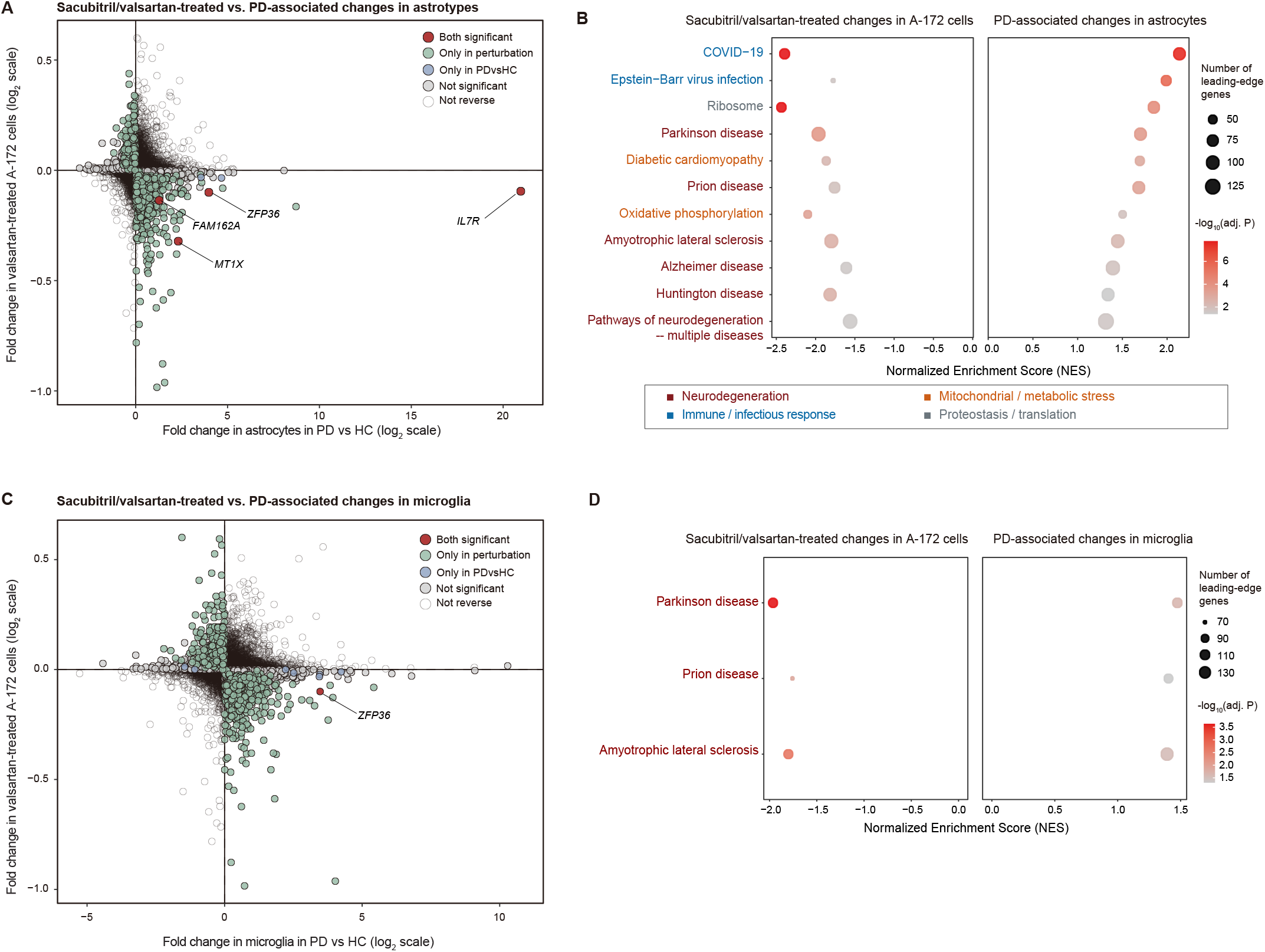
Transcriptomic analyses of sacubitril/valsartan-induced gene expression and Parkinson’s disease-associated signatures. (**A**) Gene-level scatter plot comparing log2 fold changes in PD *vs* healthy controls in astrocytes (x-axis) and sacubitril/valsartan-treated *vs* DMSO-treated A-172 cells (y-axis). Each point represents a gene. Genes with significant differential expression (FDR adjusted p < 0.05) are color-coded according to significance status in each dataset. Selected genes demonstrating bidirectional differential expression consistent with a reversal pattern are labelled. The vertical and horizontal lines indicate no differential expression. (**B**) GSEA comparing sacubitril/valsartan-treated A172 cells with DMSO-treated controls (left) and PD *vs* HC brain tissue expression profiles of astrocytes (right). Normalized enrichment scores (NES) are shown for selected KEGG pathways with FDR lower than 0.05. Circle size represents the number of leading-edge genes contributing to pathway enrichment, and colour intensity reflects -log10 adjusted *P* values. (**C**) Gene-level scatter plot comparing log2 fold changes in PD *vs* healthy controls in microglial cells (x-axis) and sacubitril/valsartan-treated *vs* DMSO-treated in A-172 cells (y-axis). Each point represents a gene. Colour coding indicates statistical significance in each dataset. (**D**) GSEA of reversed KEGG pathways comparing sacubitril/valsartan-treated A-172 cells with DMSO-treated controls (left) and PD *vs* HC brain tissue expression profiles of microglia (right). Abbreviations: FDR, false discovery rate; GSEA, gene set enrichment analysis; NES, normalized enrichment score; PD, Parkinson’s disease; HC, healthy control.

## Discussion

Massive parallel drug repurposing strategies leveraging large-scale patient data are emerging as complementary approaches to traditional preclinical pipelines in PD research.^29,30,39,40^ In this multi-stage study integrating large-scale drug-wide screening, independent cohort replication, emulated clinical trial, and exploratory cell-level transcriptomic analysis, initiation of ARBs was consistently associated with reduced risk of incident PD, with a more modest but directionally similar association observed for CCBs. Signals identified through agnostic medication screening were replicated in external PD cohorts and further evaluated in a large population patients with hypertension using an active-comparator new-user design. In the primary TTE analysis, ARB initiation was associated with approximately 30% lower hazard of PD over 10 years compared with other first-line antihypertensives, whereas CCB initiation was associated with a smaller risk reduction. The ARB association was generally consistent across many sensitivity analyses, although attenuation with a 5-year PD outcome lag and variation across active comparators highlight the potential contributions of reverse causation and residual confounding. Collectively, these findings identify a reproducible epidemiological association between ARBs and reduced PD risk but do not establish an absolutely causal or neuroprotective effect.

Several prior investigations have suggested that modulation of renin-angiotensin system (RAS) via ARB or ACEI may be associated with a lower PD risk.^5,9,12,41–43^ For example, a retrospective cohort study by Jo *et al.* using 2008-2019 healthcare claims data from Korean found that use of RAS inhibitors was significantly associated with a lower risk of PD in patients with ischemic heart disease.^12^ It reported a lower risk of PD with BBB-crossing ARBs, but not with non-BBB-crossing ARBs, suggesting that BBB permeability may modify the association between ARBs and PD risk and motivating our comparison of BBB-crossing and non-BBB-crossing ARBs. However, our exploratory analyses provided limited evidence that the observed association was attributable to BBB permeability. Moreover, individual-agent analyses relative to other first-line antihypertensives identified associations for losartan (non-BBB-crossing ARB) and valsartan (BBB-crossing ARB), but these estimates should be interpreted in the context of active comparator choice and highly unequal sample sizes across ARB agents. Together, these prior studies are either just observational, have limited sample size, or found inconsistent associations across drug classes.^9,11–13,15^ Further studies with larger numbers of ARB initiators and PD events, direct BBB-permeability-class comparisons, and more detailed characterization of BBB penetration and drug exposure are necessary to determine the observed association reflection. Similarly, CCBs have been hypothesized to influence dopaminergic neuronal function and vascular regulation, but clinical evidence has remained inconclusive.^14,44–46^ In particular, a recent study using the E3N cohort of French women reported no significant association between overall CCB use and PD in the primary 5-year-lagged analyses, while higher PD risk was observed for some non-dihydropyridine CCBs.^19^ These findings contrast with the modest inverse association observed for CCB initiation in our target trial emulation (aHR 0.88, 95% CI 0.77–0.99). Several methodological differences may contribute to this discrepancy. The E3N study compared time-varying CCB exposure with non-use in a prospective cohort of women, whereas our analysis employed an active-comparator, new-user design among individuals with primary hypertension, comparing initiation of CCBs with initiation of other first-line antihypertensive agents. Moreover, the E3N study incorporated a five-year lag between exposure assessment and PD risk, while our primary analysis began outcome follow-up after a 90-day treatment-stability period and evaluated longer outcome lags in sensitivity analyses. Importantly, our CCB association was less robust across sensitivity analyses, with attenuation toward the null in the intention-to-treat and expanded propensity-score analyses, and reduced precision in the 5-year outcome-lag analysis (**Figure 2D**). These findings, together with those of Moutard et al., suggest that associations between CCB use and PD incidence may be sensitive to exposure definitions, comparator selection, and approaches to addressing reverse causation. While we are not the first one to study the association, our study extends prior observational findings with replication cohorts and long-term TTE. The incorporation of cross-cohort replication and complementary epidemiological and transcriptomic analyses provides a more robust framework for assessing potential disease-modifying therapies. Nevertheless, the observational nature of each epidemiological analysis remains critical when interpreting consistency across datasets.

The magnitude and temporal pattern of association differed between drug classes. ARB-associated estimates were consistent across follow-up durations and sensitivity analyses, whereas CCB estimates directionally similar but attenuated over shorter follow-up windows. Notably, for ARBs, divergence between exposure groups emerged early, beginning around the first year and progressing primarily through approximately four years of follow-up, after which the curves appeared to move more nearly in parallel (**Figure 2B**). In contrast, for CCBs, little separation was observed during the first three years, with divergence becoming apparent only thereafter and continuing through later follow-up (**Figure 2C**). This pattern may suggest that the associations differ in their temporal dynamics across drug classes, with ARB-associated differences emerging earlier and stabilizing over time, whereas CCB-associated differences appear later and persist over longer follow-up. However, such observations should be interpreted cautiously, as they may also reflect residual confounding, reverse causation, or delayed ascertainment of PD rather than real time-dependent effects. This concern is particularly rational because PD has a prolonged prodromal phase, during which autonomic dysfunction and other non-motor manifestations may precede clinical diagnosis and potentially influence antihypertensive treatment selection and persistence. Therefore, we performed several analyses specifically assessing this possibility. In the TTE, additional adjustment for claims-observable characteristics related to both treatment selection and PD–including orthostatic hypotension, syncope, falls, cerebrovascular disease, tremor, constipation, REM sleep behaviour disorder, olfactory impairment, dopamine-blocking medication use, and overall medication burden–yielded an ARB estimate similar to the primary analysis. The association also remained evident with 1- and 2-year PD outcome lags but was substantially attenuated with a 5-year lag. However, these additional analyses reduce, but do not eliminate, concerns regarding reverse causation.

Moreover, given the evolving role of β-blockers in hypertension management and reported associations with increased PD risk, we excluded β-blockers from the comparator group in the sensitivity analyses and did not materially alter the protective association observed for ARBs, while estimates for CCBs remained directionally consistent but less robust.^28,31,47,48^ However, residual confounding related to comparator selection cannot be fully excluded, especially three baseline characteristics (hospitalizations, congestive heart failure, cardiac arrhythmias) remained unbalanced after IPTW. Thus, comparator choice could provide a further perspective on the observed ARB association to improve clinical interpretability. In class-specific active-comparator analyses, ARB initiation remained associated with a lower hazard of PD compared with ACEI initiation, although the magnitude was attenuated relative to the primary comparison with other first-line antihypertensives. Post-weighting balance was improved in the comparison of ARB and ACEI, with all measured covariates achieving an absolute SMD below 0.10. By contrast, no significant difference was observed between ARB and CCB initiators. Residual confounding by unmeasured clinical characteristics, however, cannot be excluded even when measured covariate balance is achieved. Furthermore, the extended follow-up period, up to a decade, provides additional context given the long prodromal phase of PD. Many prior investigations of antihypertensive medications and epilepsy have relied on relatively short follow-up periods, potentially limiting detection of long-latency neurodegenerative outcomes.^31,49^ By incorporating up to a decade of primary follow-up and additional extended observation, the present analysis provides a longer-term perspective on medication exposure and incident PD.

The transcriptomic analyses offer exploratory biological context rather than mechanistic validation. In A-172 astrocyte-like cells, transcriptional changes after sacubitril/valsartan exposure were directionally opposed to selected PD-associated gene and pathway signatures, including pathways related to inflammation, cellular stress, and neurodegeneration. Four genes involved in inflammatory signalling (*ZFP36* and *IL7R*) and oxidative stress (*FAM162A* and *MT1X*), processes increasingly recognized as central to PD pathophysiology, showed statistically significant changes in opposite directions across perturbation and PD datasets. Notably, GSEA further highlighted pathways related to the cellular response to type II interferon and antiviral immune signalling (**Supplementary Figure 7A**), suggesting that interferon-mediated host defence programs may contribute to the inflammatory landscape observed in PD. Similar transcriptional patterns were observed in microglia, suggesting consistent effects across glial cell types (**Supplementary Figure 7B**). Moreover, recent human iPSC-derived models of PD also implicated that inhibition of angiotensin receptor 1 (*AGTR1*)–the primary pharmacologic target of ARBs–protects midbrain dopaminergic neurons from degeneration and reduces α-synuclein transcription and aggregation.^50^ Complementing these experimental findings, Camacho-Meño et al. reported treatment-associated proteomic changes in circulating extracellular vesicles enriched for neuronal and glial markers from five patients with PD receiving candesartan, a BBB-penetrant ARB. These changes involved proteins associated with neuroinflammation, oxidative stress, mitochondrial function, and other PD-relevant pathways.^51^ Although these findings provide complementary human molecular evidence supporting further investigation of RAS modulation in PD, the small sample size and exploratory design preclude conclusions regarding clinical neuroprotection or disease modification. Together, these epidemiological, experimental, and molecular observations motivate further investigation of RAS modulation in PD, although they do not establish a shared mechanism or causal relationship. Interestingly, in our medication-wide screening analysis, we also observed other antihypertensive drugs targeting downstream of the renin-angiotensin-aldosterone system, such as aldosterone antagonist Spironolactone and Eplerenone, are significantly associated with reduced risk of PD (**Supplementary Figure 4)**. These studies provide complementary context but do not validate the mechanism underlying the epidemiological association observed here. The Tahoe-100M perturbation used sacubitril/valsartan, so the transcriptional response cannot be assigned specifically to valsartan or ARB activity.

From a clinical perspective, these findings should be interpreted cautiously. ARBs and CCBs are widely prescribed, generally well tolerated, and already integrated into routine cardiovascular care. The consistency of the ARB association across several epidemiological analyses supports further investigation, particularly in prospective studies designed to address long prodromal periods, treatment switching, adherence, and clinical indications for antihypertensive selection. Ultimately, randomized clinical trials or carefully designed prospective studies would be required to determine whether the observed associations reflect a causal neuroprotective effect and whether such effect is clinically meaningful.

### Limitations

This study has several limitations. First, despite the active-comparator, new-user TTE design, the analysis remains observational. Residual confounding from incompletely measured factors–including family history of PD, traumatic brain injury, smoking, alcohol use, physical activity, socioeconomic variables, frailty, renal function, and prescribing preference–cannot be excluded because of limited accessibility. Race was unavailable in the licensed MDCR data accessible for this study and therefore could not be incorporated into the TTE analysis.

Second, the primary analysis used a 90-day treatment-stability window after antihypertensive initiation, with follow-up for incident PD beginning on day 91. This approach allowed initial treatment strategy to be ascertained but conditioned inclusion on remaining PD-free and maintaining the qualifying treatment strategy during the first 90 days, potentially introducing selection bias. A sensitivity analysis beginning follow-up at treatment initiation was therefore performed. Medication exposure was based on prescription claims rather than confirmed ingestion, and adherence, treatment switching, discontinuation, and time-varying confounding could not be fully captured. Moreover, the primary analysis assessed treatment initiation rather than cumulative dose or persistent duration over time; therefore, it cannot establish whether sustained ARB exposure, which might be more biologically informative for a long-latency outcome for PD, is associated with risk. Thus, the observational emulation cannot fully reproduce treatment assignment and adherence in a randomised trial.

Third, PD outcomes were based primarily on administrative diagnostic codes and may be affected by misclassification and delayed clinical ascertainment. PD-related autonomic dysfunction, hypotension, and treatment-related blood-pressure lowering may also trigger reduction or discontinuation of antihypertensive therapy, potentially contributing to reverse causation in the case-control studies. The TTE design mitigates but cannot eliminate this concern given the prolonged prodromal phase of PD. We noticed that 20% of participants had at least one qualifying PD medication before the first PD code in MDCR, similar in MGB Biobank. Although estimates persisted with 1- and 2-year outcome lags, attenuation of the ARB estimate with a 5-year lag underscores uncertainty about prodromal PD and reverse causation; longer lags also reduce event counts and precision and can introduce selection by requiring participants to remain observable and PD-free until delayed risk entry.

Fourth, death could not be modelled as a competing event because mortality data was not sufficiently complete in MDCR; differential mortality could therefore influence PD incidence estimates.

Fifth, MDCR comprises Medicare-eligible retirees with employer-sponsored Medicare Supplemental or Medicare Advantage coverage and is not representative of all US Medicare beneficiaries or the general population. The MGB Biobank and AMP-PD cohorts likewise have distinct sampling frameworks, so cross-dataset consistency should be interpreted as complementary evidence rather than population representativeness.

Sixth, comparator choice remains important. The primary comparator pooled several antihypertensive classes, and residual heterogeneity in treatment indication may persist despite weighting and class-specific analysis. Individual-agent and BBB analyses were exploratory and had limited precision.

Finally, the transcriptomic analyses are hypothesis-generating. The perturbation involved sacubitril/valsartan, preventing attribution of the signature specifically to valsartan; A-172 is a transformed glioblastoma-derived astrocyte-like cell line; PD signatures were derived from post-mortem prefrontal cortex rather than substantia nigra or brainstem; and only four genes showed significant directional opposition at the gene level. Thus, the transcriptomic findings provide biological context but do not establish an ARB-specific mechanism or causal pathway.

## Conclusions

Initiation of angiotensin II receptor blockers and, to a lesser extent, calcium channel blockers was associated with a lower risk of incident Parkinson’s disease than initiation of other first-line antihypertensive therapies. The attenuation of the ARB estimates with a 5-year outcome lag and the remaining potential for residual confounding and reverse causation preclude causal conclusions. The results should be cautiously interpreted and do not represent an endorsement of clinical ARB use for PD prevention. Prospective studies and randomised clinical trials are needed to determine whether these associations reflect treatment effects.

## Contributors

Dong and Hu had full access to all the data in the study and take responsibility for the integrity of the data and the accuracy of the data analysis. Zhou and Xu had full access to the MarketScan data. Zhang assisted with access of the single-cell RNA-seq data. Specifically–Concept and design: Dong, Hu. Acquisition, analysis, or interpretation of data: All authors. Writing of the manuscript: Hu, Liu, Dong. Critical review of the manuscript for important intellectual content: All authors. Statistical analysis: Hu, Liu. Obtained funding: Dong, Hu. Administrative, technical, or material support: Dong, Xu. Supervision: Dong. All authors read and approved the final version of the manuscript.

## Data Sharing Statement

All analysis code is available at GitHub (https://github.com/TheDongLab/ReRx) and Zenodo (https://doi.org/10.5281/zenodo.15360497). No new primary data were collected in this study. The data and code used and generated in this study are listed in a Key Resource Table alongside their persistent identifiers at https://doi.org/10.5281/zenodo.22802220. The raw data are available upon reasonable request, following approval by the MarketScan, MGB Biobank and AMP-PD (PPMI and PDBP). An earlier version of this manuscript was posted on medRxiv on May 21, 2025, at https://doi.org/10.1101/2025.05.20.25327943.

## Data Availability

https://doi.org/10.5281/zenodo.15360497

https://doi.org/10.5281/zenodo.22802220

## Acknowledgments

Data used in the preparation of this article were obtained from the Accelerating Medicine Partnership® (AMP®) Parkinson’s Disease (AMP PD) Knowledge Platform. For up-to-date information on the study, visit https://www.amp-pd.org. The AMP® PD program is a public-private partnership managed by the Foundation for the National Institutes of Health and funded by the National Institute of Neurological Disorders and Stroke (NINDS) in partnership with the Aligning Science Across Parkinson’s (ASAP) initiative; Celgene Corporation, a subsidiary of Bristol-Myers Squibb Company; GlaxoSmithKline plc (GSK); The Michael J. Fox Foundation for Parkinson’s Research ; Pfizer Inc.; AbbVie Inc.; Sanofi US Services Inc.; and Verily Life Sciences. ACCELERATING MEDICINES PARTNERSHIP and AMP are registered service marks of the US Department of Health and Human Services. We thank the AMP PD consortium and contributing cohorts for their data contributions. The Parkinson’s Progression Markers Initiative (PPMI) is sponsored by The Michael J. Fox Foundation for Parkinson’s Research and supported by a consortium of scientific partners: 4D Pharma, Abbvie, AcureX, Allergan, Amathus Therapeutics, Aligning Science Across Parkinson’s, AskBio, Avid Radiopharmaceuticals, BIAL, BioArctic, Biogen, Biohaven, BioLegend, BlueRock Therapeutics, Bristol-Myers Squibb, Calico Labs, Capsida Biotherapeutics, Celgene, Cerevel Therapeutics, Coave Therapeutics, DaCapo Brainscience, Denali, Edmond J. Safra Foundation, Eli Lilly, Gain Therapeutics, GE HealthCare, Genentech, GSK, Golub Capital, Handl Therapeutics, Insitro, Jazz Pharmaceuticals, Johnson & Johnson Innovative Medicine, Lundbeck, Merck, Meso Scale Discovery, Mission Therapeutics, Neurocrine Biosciences, Neuron23, Neuropore, Pfizer, Piramal, Prevail Therapeutics, Roche, Sanofi, Servier, Sun Pharma Advanced Research Company, Takeda, Teva, UCB, Vanqua Bio, Verily, Voyager Therapeutics, the Weston Family Foundation and Yumanity Therapeutics. The PPMI investigators have not participated in reviewing the data analysis or content of the manuscript. For up-to-date information on the study, visit www.ppmi-info.org. The Parkinson’s Disease Biomarker Program (PDBP) consortium is supported by the National Institute of Neurological Disorders and Stroke (NINDS) at the National Institutes of Health. A full list of PDBP investigators can be found at https://pdbp.ninds.nih.gov/policy. The PDBP investigators have not participated in reviewing the data analysis or content of the manuscript. We acknowledge the Mass General Brigham (MGB) Biobank and its participants for providing samples, genomic data, and health information data. We also acknowledge the Merative™ MarketScan® Research Databases.

This research was funded in whole or in part by Aligning Science Across Parkinson’s [ASAP-000301 to X.D. and C.R.S.] and [ASAP-028376 to Y.H.] through the Michael J. Fox Foundation for Parkinson’s Research (MJFF). For the purpose of open access, the author has applied a CC BY public copyright license to all Author Accepted Manuscripts arising from this submission. Yuxuan Hu also was supported by the Exploration World Program from China Pharmaceutical University for his first-year visit to Brigham and Women’s Hospital.

During the revision of this work, we used ChatGPT (GPT-5.6 Sol) for language editing. The authors have reviewed and edited the manuscript and are fully responsible for the content of the published article.

## Research in context

### Evidence before this study

The incidence of Parkinson’s disease (PD) has been rising, yet no treatments currently exist to prevent or slow its progression. Previous observational studies have reported associations between antihypertensive medications, such as renin-angiotensin system (RAS) inhibitors and calcium channel blockers (CCBs), and reduced risk of PD. For example, a large retrospective cohort study by Jo *et al.* (2022) reported reduced PD risk associated with blood-brain barrier-penetrant angiotensin II receptor blockers (ARBs) in a Korean population with ischemic heart disease. However, prior studies were largely limited to single cohorts, conventional observational designs without explicit causal frameworks, or lacked biological interpretation. Consequently, the robustness, generalizability, and biological interpretability of these associations remain uncertain, and the extent to which such findings reflect true disease-related mechanisms versus residual bias has not been fully established.

### Added value of this study

This study adds evidence at two complementary levels. First, in the target trial emulation, initiation of ARBs was associated with a lower hazard of incident PD than initiation of other first-line antihypertensive medications; the association for CCBs was smaller and less robust. The magnitude and timing of the associations differed across drug classes, and the ARB estimate was attenuated in the 5-year outcome-lag analysis, underscoring the potential influence of the prolonged prodromal phase of PD and delayed diagnostic ascertainment. Exploratory transcriptomic analyses demonstrated directional opposition between PD-associated molecular signatures and transcriptional changes after sacubitril/valsartan exposure, but these data cannot isolate an ARB-specific effect because sacubitril has independent pharmacological activity. Second, this study integrates medication-wide screening, replication in an independent cohort, target trial emulation, and exploratory transcriptomic analyses across heterogeneous data sources. This framework strengthens the robustness of findings across datasets while retaining the limitations of observational causal inference and of cross-platform biological triangulation.

### Implications of all the available evidence

Taken together, these findings show that initiation of ARBs was associated with a lower risk of incident PD than initiation of other first-line antihypertensive medications and support further investigation of RAS modulation in PD. These findings also raise the possibility that antihypertensive selection may have implications beyond blood pressure control, particularly among individuals at elevated PD risk. The attenuation of the ARB association with a 5-year outcome lag, together with the possibility of residual confounding and reverse causation, limits causal interpretation. Randomised trials or carefully designed prospective studies are needed to determine whether these associations can translate into clinically meaningful prevention or disease modification.

